# Plasma soluble triggering receptor expressed on myeloid cells 2 is associated with depression after acute ischemic stroke

**DOI:** 10.1101/2023.04.25.23289122

**Authors:** Tiansheng Huang, Yinwei Zhu, Yaling Lu, Qi Zhang, Chongquan Fang, Zhong Ju, Jiang He, Yonghong Zhang, Tan Xu, Chongke Zhong

**Author notes:** Correspondence authors: Chongke Zhong, MD, PhD or Tan Xu, MD, PhD, Department of Epidemiology, School of Public Health and Jiangsu Key Laboratory of Preventive and Translational Medicine for Geriatric Diseases, Medical College of Soochow University, 199 Renai Road, Industrial Park District, Suzhou, Jiangsu Province 215123, China.;. **sTREM2 and Poststroke Depression**.

## Abstract

**Background:** Previous studies suggested that elevated levels of plasma soluble triggering receptor expressed on myeloid cells 2 (sTREM2) was related to increased risk of death, cardiovascular events and cognitive impairment after stroke. We aimed to prospectively investigate the association between plasma sTREM2 levels and post-stroke depression (PSD).

**Methods:** We measured plasma sTREM2 levels in 590 ischemic stroke patients from the China Antihypertensive Trial in Acute Ischemic Stroke. The 24-item Hamilton Rating Scale for Depression was used to assess depression at 3 months after ischemic stroke onset, and PSD was defined as a score of ≥8. Logistic regression analysis was performed to evaluate the risk of PSD associated with plasma sTREM2 levels, and net reclassification index (NRI) and integrated discrimination improvement (IDI) were calculated to assess the predictive value of sTREM2.

**Results:** Of the 590 participants, 229 (38.8%) patients experienced PSD. The risk of PSD elevated significantly with plasma sTREM2 levels (*P* for trend=0.034). After adjusting for several covariates, the odds ratio for the highest quartile of sTREM2 compared with the lowest quartile was 2.41 (95% CI=1.35-4.31) for PSD. Multiple adjusted spline regression analysis further confirmed the linear dose-response relationship between sTREM2 levels and PSD (*P* for linearity=0.024). The addition of sTREM2 to a conventional model notably improved the risk prediction for PSD (category-free NRI=21.50%, 95% CI=5.92%-37.07%, *P*=0.011; IDI=1.43%, 95% CI=0.45%-2.42%, *P*=0.005).

**Conclusions:** The present study demonstrated that elevated plasma sTREM2 levels were associated with increased risk of PSD, suggesting that sTREM2 may be a promising prognostic biomarker for PSD.

**Registration:** URL: https://www.clinicaltrials.gov; Unique identifier: NCT01840072.

## Background

Stroke is the second leading cause of death worldwide and a major cause of disability. Due to the aging of the population, the incidence of stroke continues to rise globally, with ischemic stroke accounting for almost 87 percent^1^. Although the age-standardized incidence and mortality of stroke in China has declined since 1990, the disease burden of stroke remains significant^2^. Post-stroke depression (PSD) is the most frequent and burdensome neuropsychiatric complication after stroke, and the prevalence of PSD is estimated to be between 30% and 37% in a meta-analysis^3, 4^. PSD is associated with disability, social function defects, and increased stroke mortality^5^. The identification of effective biomarkers for PSD is urgently required to provide valuable predictive information and improve clinical care.

Microglia are the main immune cells and the first line of defense against injury in the central nervous system^6^. However, when injury-related molecular patterns are overactivated after stroke, microglia produce a large number of proinflammatory cytokines, further damage neural cells and the blood-brain barrier, and affect vascular remodeling^7^. Triggering receptor expressed on myeloid cells 2 (TREM2), a receptor expressed on the surface of microglia, are significantly associated with microglial activity^8^. The ectodomain of TREM2 is processed by a disintegrin and metalloproteinase, resulting in the release of a soluble form of TREM2 (sTREM2). sTREM2 can promote the survival of microglia and stimulate the production of inflammatory cytokines, and contribute to the development of neurodegenerative disorders^9^. Previous studies demonstrated that elevated plasma sTREM2 levels were associated with increased risk of death, cardiovascular events, and cognitive impairment after ischemic stroke^10, 11^. However, population-based evidence on the relationship between sTREM2 and PSD are sparse.

Recently, cerebrospinal fluid (CSF) sTREM2 were found to be associated with the markers of cerebrovascular injury and neurodegeneration, and subjects with elevated levels of CSF sTREM2 were more likely to acquire delirium^12, 13^. Furthermore, a cohort study in Chinese population over 40 years revealed that minimal depressant symptoms were associated with CSF sTREM2 levels^14^. We hypothesized that circulating sTREM2 would offer promising prognostic significance for PSD among patients with acute ischemic stroke.

To further extend the current knowledge on the potential predictive role of sTREM2 for PSD, we prospectively explored the relationship between plasma sTREM2 and PSD using data from the China Antihypertensive Trial in Acute Ischemic Stroke (CATIS).

## Methods

The data that support the findings of this study are available from the corresponding author upon reasonable request.

### Trial design and participants

The participants in this study were from the CATIS trial conducted in 26 hospitals, and the design details have been described previously^15^. In brief, the CATIS trial was a multicenter, single-blind, blind end-point randomized clinical trial. The inclusion criteria were as follows: (1) age ≥22 years; (2) first-ever ischemic stroke diagnosed by neurologist using computed tomography or magnetic resonance imaging; (3) time from onset to admission within 48 h; (4) systolic blood pressure (BP) between 140 and 220 mmHg. The exclusion criteria were as follows: (1) BP ≥220/120 mmHg; (2) treated with intravenous thrombolytic therapy; (3) severe heart failure, acute myocardial infarction or unstable angina pectoris, atrial fibrillation, aortic dissection, cerebrovascular stenosis, resistant hypertension, or deep coma. Finally, 4071 patients with elevated BP were recruited in the CATIS trial from August 2009 and May 2013. The present study was based on a preplanned ancillary study of CATIS trial, which was designed to investigate whether early BP reduction could reduce cognitive impairment at 3 months in patients with acute ischemic stroke^16^. In the ancillary study, 7 of 26 hospitals were randomly selected and 660 participants were consecutively recruited for neuropsychiatric status assessment from August 2009 to November 2012. At the 3-month visit, 15 patients were lost to follow-up and 7 died. Of the 638 participants who eventually completed the cognitive function tests, 590 patients were eligible for this analysis because some refused to provide blood samples or the collected samples were hemolyzed during storage or transport.

The study conforms to the World Medical Association Declaration of Helsinki, and was approved by the institutional review boards at Soochow University in China and Tulane University in the United States, as well as ethical committees at the participating hospitals. Written consent was obtained from all study participants or their immediate family members.

### Assessment of sTREM2 concentration and potential covariates

Blood samples were collected after at least 8 h of fasting and within 24 h of hospital admission, and were separated and frozen at −80°C in the Central Laboratory of the School of Public Health at Soochow University. Circulating sTREM2 and galectin-3 concentrations were measured centrally at Soochow University using commercially available immunoassays (R&D Systems). The intra-assay and inter-assay coefficients of variations for the two biomarkers were below 3.6% and 6.9%, respectively. Measurements were performed by laboratory technicians who were blind to the clinical characteristics and outcomes of the participants.

Baseline data on demographic characteristics, clinical characteristics, medical history, and medication history were collected by trained interviewers using standard questionnaires at enrollment. Blood lipid, glucose and other clinical laboratory measurements were performed for all enrolled patients in each participating hospital at admission. BP was measured by trained nurses according to a standard protocol adapted from procedures recommended by the American Heart Association and stroke severity was assessed by neurologists using the National Institutes of Health Stroke Scale (NIHSS) at admission^17, 18^. The subtype of ischemic stroke was classified as thrombotic (large artery atherosclerosis), embolic (cardiac embolism) and lacunar (small artery occlusive lacunar) according to clinical features and imaging data^19^.

### Assessment of outcomes

The depression status at 3 months after acute ischemic stroke was assessed by neurologists using the Hamilton Rating Scale for Depression (HRSD-24)^20, 21^. The HRSD-24 scale has been translated into Chinese and widely used to measure the severity of depression with good reliability and validity^22^. The cut-off point for diagnosing depression is recommended to be 8^23^. According to the severity of depression, participants were categorized as follows: no depression (0 to 7 points), mild depression (8 to 19 points), and severe depression (20 points and above)^24^.

### Statistical analysis

All participants were divided into four groups according to sTREM2 level quartiles. Baseline characteristics were displayed as mean with standard deviation (SD), median with interquartile range (IQR), or frequency with percentage, depending on the distribution. The generalized linear regression analysis and Cochran-Armitage trend χ^2^ test were used to test the interquartile trends for continuous variables and categorical variables, respectively. Logistic regression analyses were performed to estimate the risk of PSD associated with sTREM2 quartiles by calculating odds ratios (ORs) and 95% confidence intervals (CIs), as well as the risk for each SD increase in logarithmic transformation of sTREM2 levels. The impact of sTREM2 on PSD severity was analyzed using the ordinal logistic regression model. Potential covariates including age, sex, educational level, current smoking, alcohol drinking, time from onset to randomization, blood glucose, platelet counts, creatinine, homocysteine, galectin-3, total cholesterol, diastolic BP, baseline NIHSS score, body mass index (BMI), randomized treatment, family history of stroke, medical history (diabetes mellitus, hypertension and hyperlipidemia), use of lipid-lowering medications, use of antihypertensive medications and ischemic stroke subtype were adjusted in multivariable analyses. Potential covariates for PSD were selected according to previous reports^10, 25^. The linear trend of OR across the sTREM2 quartiles was tested using the median within each quartile in the logistic regression model as a predictor.

Spline regression model was performed to explore the shape of the association between the sTREM2 level and PSD, fitting a restricted cubic spline with four sections (5th, 35th, 65th and 95th percentiles)^26^. Moreover, we performed subgroup analyses to explore the potential effect modification by risk factors including age, sex, education, baseline diastolic BP, BMI, smoking and alcohol consumption, baseline NIHSS score, family history of stroke, and subtype of ischemic stroke. The likelihood ratio test of the interaction term model was used to test the interaction between sTREM2 and the factors mentioned above. In addition, we tested the performance of models with sTREM2 to predict the risk of PSD. First, net reclassification index (NRI) and integrated discrimination improvement (IDI) were calculated to assess the improvement in risk classification after the addition of sTREM2 to the basic model with conventional risk factors^27^. Second, the C-statistics were estimated to compare the risk discrimination for PSD between the basic model with and without plasma sTREM2 levels. Third, the Hosmer-Lemeshow χ^2^ statistics were used to evaluate the calibration of models. Markov chain Monte Carlo method was used to perform multiple imputations on missing data. All *P*-values were double-tailed and a significance level of 0.05 was adopted. SAS statistical software was used for statistical analysis (version 9.4; SAS Institute).

## Results

### Baseline characteristics

Among the 590 patients, the mean age was 59.8±10.4 years, and 420 (71.2%) were men. The median level of sTREM2 was 404.7 pg/ mL (IQR: 265.1 to 602.5 pg/ mL). As shown in **Table 1**, compared with participants with lower sTREM2 levels, participants with higher sTREM2 levels were more likely to be older and female, to have higher plasma galectin-3 concentrations, lower baseline diastolic BP, BMI, education levels and prevalence of alcohol drinking (*P* for trend<0.05).

**Table 1.**
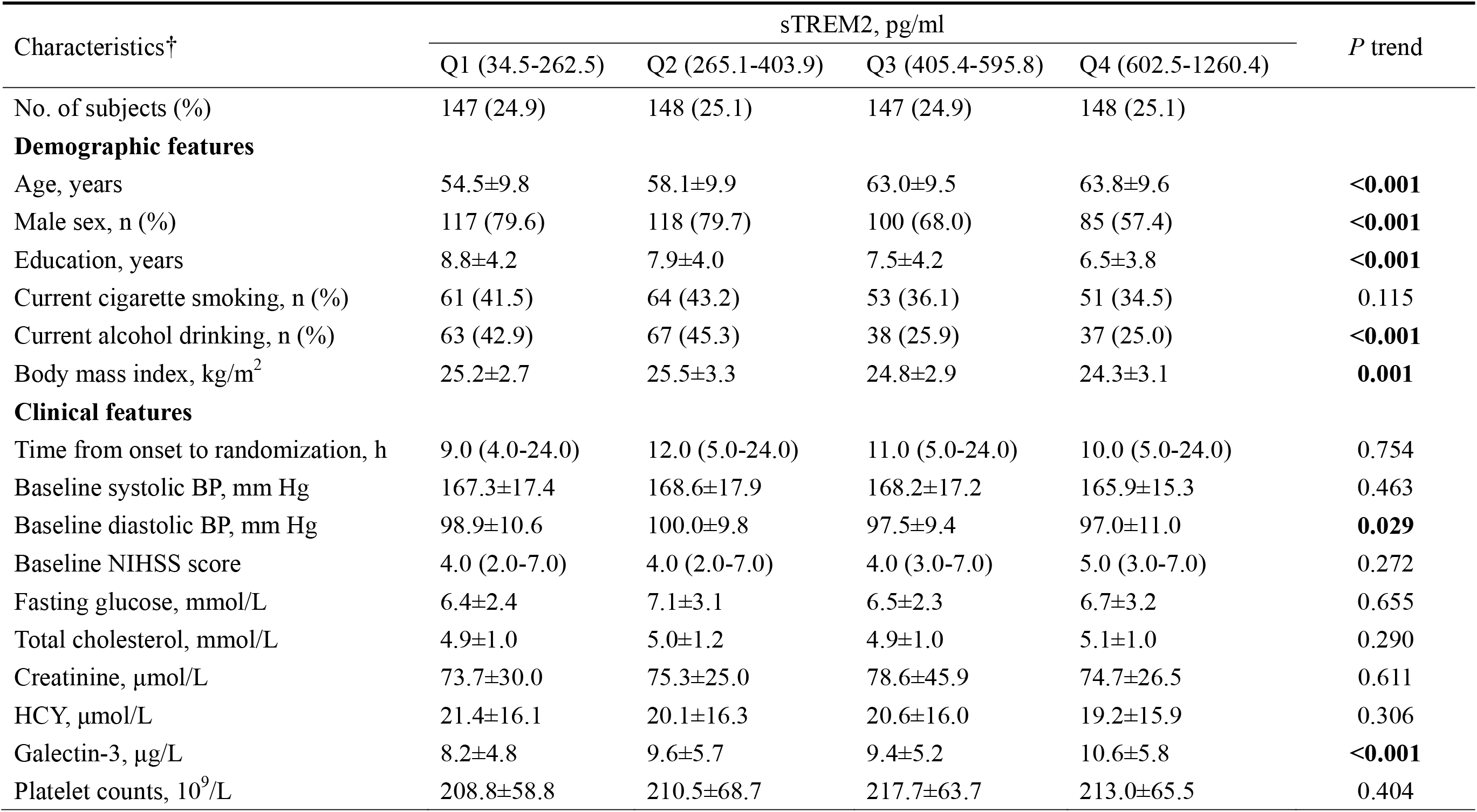
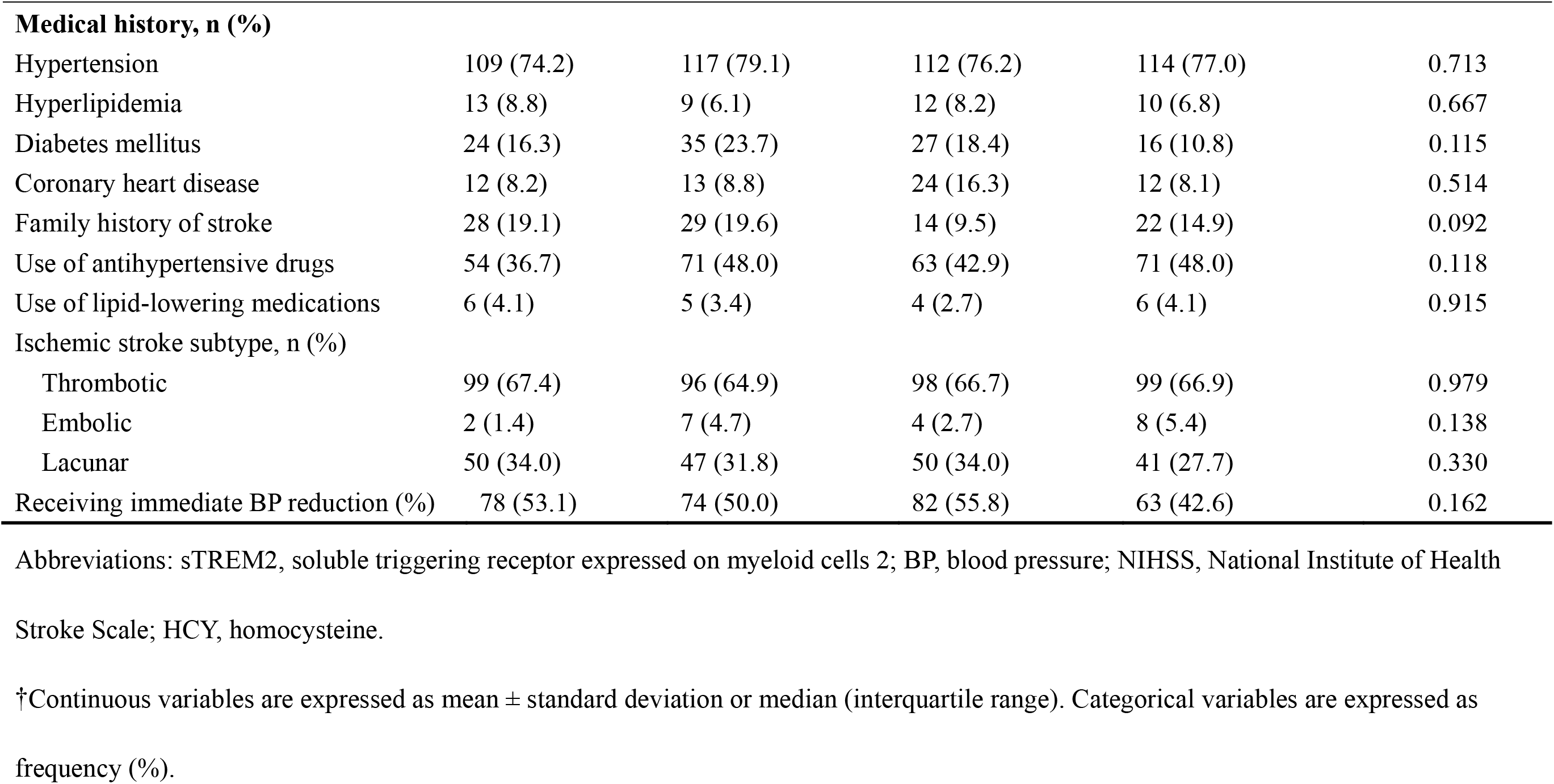
Baseline Characteristics of Participants According to Plasma soluble TREM2 quartiles.

| Characteristics<sup>†</sup> | sTREM2, pg/ml |  |  |  | *P* trend |
| --- | --- | --- | --- | --- | --- |
|  | Q1 (34.5-262.5) | Q2 (265.1-403.9) | Q3 (405.4-595.8) | Q4 (602.5-1260.4) |  |
| No. of subjects (%) | 147 (24.9) | 148 (25.1) | 147 (24.9) | 148 (25.1) |  |
| **Demographic features** |  |  |  |  |  |
| Age, years | 54.5±9.8 | 58.1±9.9 | 63.0±9.5 | 63.8±9.6 | **<0.001** |
| Male sex, n (%) | 117 (79.6) | 118 (79.7) | 100 (68.0) | 85 (57.4) | **<0.001** |
| Education, years | 8.8±4.2 | 7.9±4.0 | 7.5±4.2 | 6.5±3.8 | **<0.001** |
| Current cigarette smoking, n (%) | 61 (41.5) | 64 (43.2) | 53 (36.1) | 51 (34.5) | 0.115 |
| Current alcohol drinking, n (%) | 63 (42.9) | 67 (45.3) | 38 (25.9) | 37 (25.0) | **<0.001** |
| Body mass index, kg/m<sup>2</sup> | 25.2±2.7 | 25.5±3.3 | 24.8±2.9 | 24.3±3.1 | **0.001** |
| **Clinical features** |  |  |  |  |  |
| Time from onset to randomization, h | 9.0 (4.0-24.0) | 12.0 (5.0-24.0) | 11.0 (5.0-24.0) | 10.0 (5.0-24.0) | 0.754 |
| Baseline systolic BP, mm Hg | 167.3±17.4 | 168.6±17.9 | 168.2±17.2 | 165.9±15.3 | 0.463 |
| Baseline diastolic BP, mm Hg | 98.9±10.6 | 100.0±9.8 | 97.5±9.4 | 97.0±11.0 | **0.029** |
| Baseline NIHSS score | 4.0 (2.0-7.0) | 4.0 (2.0-7.0) | 4.0 (3.0-7.0) | 5.0 (3.0-7.0) | 0.272 |
| Fasting glucose, mmol/L | 6.4±2.4 | 7.1±3.1 | 6.5±2.3 | 6.7±3.2 | 0.655 |
| Total cholesterol, mmol/L | 4.9±1.0 | 5.0±1.2 | 4.9±1.0 | 5.1±1.0 | 0.290 |
| Creatinine, µmol/L | 73.7±30.0 | 75.3±25.0 | 78.6±45.9 | 74.7±26.5 | 0.611 |
| HCY, µmol/L | 21.4±16.1 | 20.1±16.3 | 20.6±16.0 | 19.2±15.9 | 0.306 |
| Galectin-3, µg/L | 8.2±4.8 | 9.6±5.7 | 9.4±5.2 | 10.6±5.8 | **<0.001** |
| Platelet counts, 10<sup>9</sup>/L | 208.8±58.8 | 210.5±68.7 | 217.7±63.7 | 213.0±65.5 | 0.404 |
| **Medical history, n (%)** |  |  |  |  |  |
| Hypertension | 109 (74.2) | 117 (79.1) | 112 (76.2) | 114 (77.0) | 0.713 |
| Hyperlipidemia | 13 (8.8) | 9 (6.1) | 12 (8.2) | 10 (6.8) | 0.667 |
| Diabetes mellitus | 24 (16.3) | 35 (23.7) | 27 (18.4) | 16 (10.8) | 0.115 |
| Coronary heart disease | 12 (8.2) | 13 (8.8) | 24 (16.3) | 12 (8.1) | 0.514 |
| Family history of stroke | 28 (19.1) | 29 (19.6) | 14 (9.5) | 22 (14.9) | 0.092 |
| Use of antihypertensive drugs | 54 (36.7) | 71 (48.0) | 63 (42.9) | 71 (48.0) | 0.118 |
| Use of lipid-lowering medications | 6 (4.1) | 5 (3.4) | 4 (2.7) | 6 (4.1) | 0.915 |
| Ischemic stroke subtype, n (%) |  |  |  |  |  |
| Thrombotic | 99 (67.4) | 96 (64.9) | 98 (66.7) | 99 (66.9) | 0.979 |
| Embolic | 2 (1.4) | 7 (4.7) | 4 (2.7) | 8 (5.4) | 0.138 |
| Lacunar | 50 (34.0) | 47 (31.8) | 50 (34.0) | 41 (27.7) | 0.330 |
| Receiving immediate BP reduction (%) | 78 (53.1) | 74 (50.0) | 82 (55.8) | 63 (42.6) | 0.162 |

Abbreviations: sTREM2, soluble triggering receptor expressed on myeloid cells 2; BP, blood pressure; NIHSS, National Institute of Health
Stroke Scale; HCY, homocysteine.
†Continuous variables are expressed as mean ± standard deviation or median (interquartile range). Categorical variables are expressed as frequency (%).

### Association between sTREM2 and PSD

The median (IQR) score of the HRSD-24 was 5 (1-12). A total of 229 patients (38.8%) experienced depression at 3-month follow-up. The risk of PSD elevated significantly with plasma sTREM2 levels (*P* for trend=0.034; **Table 2**). After adjusting for potential covariates, including age, sex, education, smoke, drink, baseline NIHSS score and other confounding factors, the adjusted OR (95% CI) of the second, third and highest quartiles were 1.86 (95% CI=1.08-3.22), 1.94 (95% CI=1.11-3.37) and 2.41 (95% CI=1.35-4.31), respectively, compared with the lowest sTREM2 quartile. Per SD increase in logarithmic sTREM2 was associated with a 26% (95% CI=2%-55%) increase in PSD risk. Multiple adjusted spline regression analysis further confirmed the linear dose-response relationship between sTREM2 level and PSD (*P*=0.024; **Figure 1**). Subgroup analysis revealed that the previously mentioned important covariates did not change the relationship between sTREM2 and PSD (*P* >0.05 for interaction; **Table 3**). The ordinal logistic regression model indicated that increased sTREM2 was significantly associated with depression severity after adjusting for covariates (OR=2.19; 95% CI=1.25-3.83; *P* for trend=0.010, when the highest and the lowest quartile were compared).

**Figure 1.**
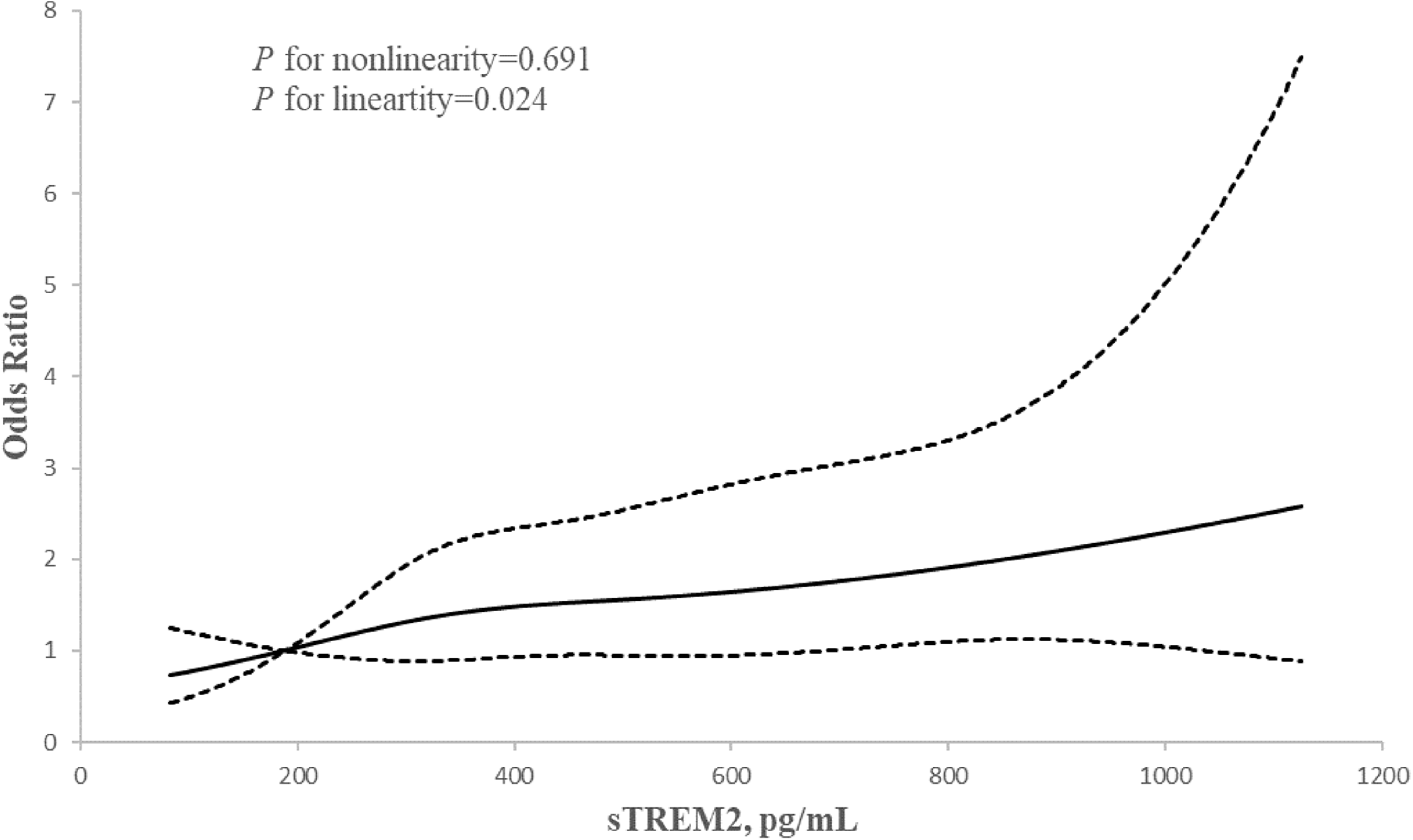
Association of sTREM2 with risk of depression after acute ischemic stroke. Odds ratios and 95% confidence intervals derived from restricted cubic spline regression, with knots placed at the 5th, 35th, 65th, and 95th percentiles of the distribution of sTREM2. The reference point for sTREM2 is the midpoint (189.0 pg/ml) of the reference group from categorical analysis. Odds ratios were adjusted for the same variables as multivariable adjusted model in Table 2.

**Table 2.** ORs and 95% CIs for the risk of post-stroke depression according to sTREM2 quartiles.

|  | sTREM2, pg/ml |  |  |  | *P* value<br>for trend | Each SD increase in<br>logarithm sTREM2 |
| --- | --- | --- | --- | --- | --- | --- |
|  | Q1 (34.5-262.5) | Q2 (265.1-403.9) | Q3 (405.4-595.8) | Q4 (602.5-1260.4) |  |  |
| Median | 189.0 | 325.8 | 487.6 | 801.6 | - | - |
| **Poststroke depression, HRSD score≥8** |  |  |  |  |  |  |
| Cases, n (%) | 44 (29.93) | 60 (40.54) | 63 (42.86) | 62 (41.89) | **0.034** | - |
| Unadjusted | 1.00 | 1.60 (0.99-2.58) | 1.76 (1.09-2.84) | 1.69 (1.04-2.73) | **0.034** | 1.15 (0.97-1.36) |
| Multivariable adjusted | 1.00 | 1.86 (1.08-3.22) | 1.94 (1.11-3.37) | 2.41 (1.35-4.31) | **0.005** | 1.26 (1.02-1.55) |
| **Poststroke depression severity** |  |  |  |  |  |  |
| Unadjusted | 1.00 | 1.63 (1.01-2.63) | 1.78 (1.11-2.87) | 1.66 (1.03-2.68) | **0.040** | 1.14 (0.97-1.35) |
| Multivariable adjusted | 1.00 | 1.83 (1.08-3.09) | 1.82 (1.07-3.11) | 2.19 (1.25-3.83) | **0.010** | 1.22 (1.00-1.49) |

Multivariable adjusted model: adjusted for age, sex, educational level, current smoking, alcohol drinking, time from onset to randomization, blood glucose, body mass index, galectin-3, platelet counts, homocysteine, creatinine, total cholesterol, diastolic blood pressure, baseline National Institutes of Health Stroke Scale score, randomized treatment, medical history (diabetes mellitus, hypertension and hyperlipidemia), family history of stroke, use of lipid-lowering medications, use of antihypertensive medications and ischemic stroke subtype.
Abbreviations: sTREM2, soluble triggering receptor expressed on myeloid cells 2; SD, standard deviation; HRSD, Hamilton Rating Scale for Depression; CI, confidence interval; and OR, odds ratio.

**Table 3.** Subgroup analyses of the association between plasma sTREM2 and post-stroke depression.

| Subgroup | sTREM2, pg/ml |  |  |  | *P* trend | *P* interaction |
| --- | --- | --- | --- | --- | --- | --- |
|  | Q1 (34.5-262.5) | Q2 (265.1-403.9) | Q3 (405.4-595.8) | Q4 (602.5-1260.4) |  |  |
| Stroke onset age, years |  |  |  |  |  | 0.40 |
| <65 | 1.00 | 1.60 (0.84-3.07) | 2.91 (1.48-5.71) | 2.33 (1.11-4.91) | **<0.01** |  |
| ≥65 | 1.00 | 2.72 (0.82-9.04) | 1.15 (0.38-3.53) | 3.58 (1.10-11.62) | 0.12 |  |
| Sex |  |  |  |  |  | 0.94 |
| Male | 1.00 | 1.74 (0.95-3.21) | 1.78 (0.93-3.38) | 2.54 (1.27-5.10) | **0.01** |  |
| Female | 1.00 | 1.94 (0.46-8.09) | 2.56 (0.72-9.16) | 2.33 (0.65-8.29) | 0.23 |  |
| Education, years |  |  |  |  |  | 0.83 |
| <8 | 1.00 | 2.83 (1.11-7.26) | 1.47 (0.57-3.82) | 2.83 (1.11-7.20) | 0.11 |  |
| ≥8 | 1.00 | 1.56 (0.72-3.38) | 2.52 (1.15-5.53) | 2.12 (0.87-5.19) | **0.03** |  |
| Baseline diastolic BP |  |  |  |  |  | 0.61 |
| <100 | 1.00 | 1.14 (0.50-2.60) | 1.84 (0.84-4.03) | 2.16 (0.96-4.86) | **0.04** |  |
| ≥100 | 1.00 | 3.27 (1.42-7.53) | 1.96 (0.80-4.76) | 2.73 (1.05-7.10) | 0.15 |  |
| Body mass index, kg/m<sup>2</sup> |  |  |  |  |  | 0.54 |
| <24 | 1.00 | 2.21 (0.75-6.56) | 2.04 (0.70-5.96) | 2.53 (0.88-7.30) | 0.14 |  |
| ≥24 | 1.00 | 2.00 (1.02-3.92) | 1.89 (0.95-3.77) | 2.78 (1.30-5.92) | **0.01** |  |
| Cigarette smoking |  |  |  |  |  | 0.09 |
| No | 1.00 | 1.70 (0.81-3.57) | 1.09 (0.52-2.29) | 1.72 (0.80-3.70) | 0.33 |  |
| Yes | 1.00 | 1.90 (0.73-4.92) | 4.81 (1.81-12.80) | 4.83 (1.63-14.28) | **<0.01** |  |
| Alcohol consumption |  |  |  |  |  | 0.27 |
| No | 1.00 | 1.15 (0.52-2.54) | 1.27 (0.62-2.60) | 1.63 (0.76-3.50) | 0.19 |  |
| Yes | 1.00 | 2.82 (1.17-6.77) | 2.96 (1.05-8.34) | 3.65 (1.21-10.98) | **0.02** |  |
| Admission NIHSS score |  |  |  |  |  | 0.50 |
| <4 | 1.00 | 2.23 (0.85-5.88) | 1.68 (0.61-4.60) | 1.40 (0.46-4.23) | 0.61 |  |
| ≥4 | 1.00 | 1.99 (0.96-4.13) | 2.01 (0.99-4.08) | 2.85 (1.36-5.98) | **<0.01** |  |
| Family history of stroke |  |  |  |  |  | 0.76 |
| No | 1.00 | 1.42 (0.78-2.60) | 1.84 (1.01-3.33) | 2.30 (1.22-4.33) | **<0.01** |  |
| Yes | 1.00 | 3.07 (0.49-19.16) | 1.29 (0.14-11.67) | 1.95 (0.23-16.31) | 0.77 |  |
| Ischemic stroke subtype |  |  |  |  |  | 0.10 |
| Thrombotic | 1.00 | 2.69 (1.29-5.64) | 3.37 (1.58-7.21) | 4.04 (1.83-8.90) | **<0.01** |  |
| Non-Thrombotic | 1.00 | 1.03 (0.37-2.86) | 0.66 (0.23-1.83) | 1.13 (0.39-3.28) | 0.96 |  |

Odds ratios (ORs) were calculated after adjustment for the same variables as adjusted model in Table 2, except for the stratified variable.
Abbreviations: sTREM2, soluble triggering receptor expressed on myeloid cells 2; BP, blood pressure; NIHSS, National Institute of Health
Stroke Scale.

We further investigated the incremental predictive value of sTREM2 for PSD beyond traditional risk factors (**Table 4**). Adding sTREM2 to the conventional risk factors model notably improved risk reclassification for PSD, shown by an increase in category-free NRI of 21.50% (95% CI=5.92%-37.07%; *P*=0.011) and IDI of 1.43% (95% CI=0.45%-2.42%; *P*=0.005). Furthermore, plasma sTREM2 marginally increased the C-statistic from 0.732 (95%CI=0.690-0.774) to 0.745 (95%CI=0.704-0.786; *P* for difference=0.06). The Hosmer-Lemeshow test showed that the model calibration was adequate after the addition of sTREM2 to the basic model with traditional risk factors (*P*=0.96).

**Table 4.** Reclassification Statistics (95% CI) for post-stroke depression by plasma sTREM2 among patients with acute ischemic stroke.

|  | C statistic |  | Calibration<br>statistic |  | NRI (category-free) |  | IDI |  |
| --- | --- | --- | --- | --- | --- | --- | --- | --- |
| | Estimate (95% CI) | *P*<br>value | $\chi^2$ | *P*<br>value | Estimate (95% CI), % | *P*<br>value | Estimate (95% CI), % | *P*<br>value |
| Conventional model | 0.732 (0.690 to 0.774) |  | 5.94 | 0.65 | Reference |  | Reference |  |
| Conventional model +<br>sTREM2<sup>†</sup> | 0.745 (0.704 to 0.786) | 0.06 | 2.55 | 0.96 | 21.50 (5.92 to 37.07) | 0.011 | 1.43 (0.45 to 2.42) | 0.005 |

Conventional model included age, sex, educational level, current smoking, alcohol drinking, time from onset to randomization, blood glucose, body mass index, galectin-3, platelet counts, homocysteine, creatinine, total cholesterol, diastolic blood pressure, baseline National Institutes of Health Stroke Scale score, randomized treatment, medical history (diabetes mellitus, hypertension and hyperlipidemia), family history of stroke, use of lipid-lowering medications, use of antihypertensive medications and ischemic stroke subtype.
Abbreviations: CI, confidence interval; sTREM2, soluble triggering receptor expressed on myeloid cells 2; IDI, integrated discrimination index; and NRI, net reclassification improvement.
<sup>†</sup>quartiles.

## Discussion

To our knowledge, this study is the first to explore the association between plasma sTREM2 levels and 3-month PSD among Chinese ischemic stroke patients. We demonstrated that higher plasma sTREM2 levels were associated with an increased risk of PSD after adjusting for confounders, and there was a linear dose-response relationship between sTREM2 levels and PSD. Furthermore, the addition of sTREM2 to the conventional risk factors model significantly improved the risk prediction for PSD. These results indicated that plasma sTREM2 could play an important role in predicting PSD.

TREM2 is a congenital immune receptor localized on microglia. It belongs to the immunoglobulin and lectin like superfamily and is involved in microglia survival, inflammatory response, and induction of phagocytosis^8, 28^. A population-based case-control study reported significantly elevated TREM2 mRNA expression in peripheral leukocytes of patients with schizophrenia^29^. Experimental study found that rats injected intraperitoneally with lipopolysaccharide exhibited depression like behavior and increased TREM2 expression in the hippocampus and prefrontal cortex compared with the control group^30^. The ectodomain of TREM2 is processed by a disintegrin and metalloprotease, resulting in the release of a soluble form of TREM2, which can be detected in peripheral blood and CSF^31^. sTREM2 has been shown to be associated with the development and progression of neurological diseases, such as Alzheimer’s disease, obstructive sleep apnea and sepsis related encephalopathy^13, 32, 33^. In a cross-sectional study of 94 patients with obstructive sleep apnea, serum sTREM2 levels were higher in patients with mild cognitive impairment than in those without^33^. Recently, in a previous population-based study, our team found that the risk of post-stroke cognitive impairment increased significantly with the increase of plasma sTREM2 levels among patients with acute ischemic stroke^11^.

PSD is another common neuropsychiatric complication after ischemic stroke, however, very limited studies have reported the relationship between sTREM2 and PSD. Henjum et al. conducted a cohort study of 59 patients with hip fractures and found that elevated CSF sTREM2 levels were associated with the risk of delirium^12^. Another cohort study involving 796 cognitively unimpaired Han Chinese aged 40-90 reported that minimal depressive symptoms were associated with CSF sTREM2 levels, and the sTREM2-amyloid pathway partially mediated the impact of mild depressive symptoms on cognition^14^. Therefore, plasma sTREM2 might be a valuable predictor of depression. Our results confirmed this hypothesis and extended the predictive value of plasma sTREM2 to the ischemic stroke population. Plasma sTREM2 was independently associated with PSD risk and had incremental prognostic value beyond traditional risk factors, suggesting that sTREM2 may be a promising prognostic biomarker for PSD.

The mechanisms of the relationship between sTREM2 and PSD remain unclear, but several possible pathophysiological pathways have been proposed. First, sTREM2 has been reported to promote the survival of microglia, stimulate the production of inflammatory cytokines, and further damage neural cells, blood-brain barrier and disrupt cerebrovascular remodeling after brain injury^9, 34^. Impaired hippocampal neurogenesis and inhibition of neuroplasticity induced by activation of microglia are considered to be important mechanisms of depression and targets of antidepressants^35^. In addition, TREM2 can induce the activation of M2 phenotypes microglia after nerve injury to promote the phagocytosis of apoptotic neurons and inhibit the expression of pro-inflammatory molecules, thereby alleviating inflammation and promoting tissue repair. Interestingly, sTREM2 was reported to inhibit the neuroprotective function of TREM2 by competitively binding to TREM2 ligands^36^. More research is needed to explain the mechanisms underlying the association between sTREM2 and PSD.

The present study is a prospective cohort study from the CATIS trial with standardized protocols and strict quality control procedures for data collection and outcome assessment. Moreover, comprehensive information about potential confounders was collected at baseline. These strengths contribute to providing a valid assessment of the association between sTREM2 and PSD. In addition, previous studies reported the predictive role of CSF sTREM2 levels for depression^14^. However, CSF collection is invasive, so the prediction of PSD by plasma sTREM2 in this study is more suitable for clinical use due to noninvasiveness and convenience. Of note, several limitations needed to be interpreted. First, the subjects in our study were only from a randomized subsample of the CATIS trial, excluding ischemic stroke patients with BP ≥220/120 mmHg or receiving intravenous thrombolytic treatment. Therefore, selection bias was inevitable. However, the proportion of patients with BP ≥220/120 mmHg or receiving intravenous thrombolytic treatment is low in China^37^. Further large prospective cohort studies in different populations are warranted to verify our findings. Second, in this study, plasma sTREM2 levels were measured only once during the acute phase of ischemic stroke, which limited our study of its dynamic change over time and its implications for clinical stroke outcomes. Third, data on baseline psychiatric history were not collected, which could confound our findings. However, it was reported that the NIHSS score had a subset of cognitive assessments, and that depression was closely correlated with sex, age, stroke severity, history of diabetes mellitus, and other baseline characteristics^38, 39^. We adjusted for these covariates in the analysis to reduce the potential bias of baseline depression on the results. Final, the present study was observational. Although we adjusted for multiple covariates, the causality of plasma sTREM2 and PSD could still not be established due to the presence of residual confounders such as TREM2 variants and brain imaging biomarkers.

In conclusion, our results suggested that elevated plasma sTREM2 levels in the acute phase of ischemic stroke were associated with the risk of PSD, independently of traditional risk factors.

## Acknowledgements

We thank the study participants and their relatives and the clinical staff at all participating hospitals for their support and contribution to this project.

## Supplemental Material

Tables S1-S3

## Nonstandard Abbreviations and Acronyms

CATIS: China Antihypertensive Trial in Acute Ischemic Stroke
NIHSS: National Institutes of Health Stroke Scale
sTREM2: soluble triggering receptor expressed on myeloid cells 2

